# Polygenic risk associations with developmental and mental health outcomes in childhood and adolescence: A systematic review

**DOI:** 10.1101/2023.03.31.23287877

**Authors:** L.B. Moyakhe, S. Dalvie, M.S. Mufford, D.J. Stein, N. Koen

## Abstract

**Background:** Neurodevelopmental and mental health disorders in childhood constitute an emerging global concern, with adverse sequelae which span children’s physical, psychological and social well-being. The aetiology of these disorders is likely complex, multifactorial and polygenic. Polygenic risk scores (PRS), an estimate of an individual’s genetic liability toward a disorder, have been increasingly used in psychiatric research to explore genetic associations with disorders of interest. However, limited work delineates polygenic associations with development and mental health in childhood populations.

We aimed to systematically review existing literature on associations between genetic risk (as measured by PRS) and neurodevelopmental and mental health outcomes in childhood and adolescence.

**Methods:** Following the recommended Preferred Reporting Items for Meta-Analyses (PRISMA) guidelines, databases were searched using key search terms. The search commenced in March 2021 and concluded in June 2021. The studies eligible for inclusion were full-text articles investigating polygenic risk associations with neurodevelopmental and/or mental health outcomes in childhood or adolescence.

**Results:** Fourteen studies were eligible for inclusion in this systematic review. The association between higher PRS for attention-deficit/hyperactivity disorder (ADHD) and adverse developmental/mental health outcomes in childhood and adolescence was reported by five studies. Additionally, associations between PRS for bipolar disorder or major depressive disorder and adverse outcomes of interest were also described by two studies; and two studies highlighted associations between schizophrenia PRS and mental health disorders in childhood. The remaining studies highlighted shared polygenic contributions between and within NDDs and mental health disorders in children.

**Conclusion:** The findings of this systematic review suggest that PRS for neurodevelopmental and mental health disorders may associate with adverse neurodevelopmental and mental health outcomes from early childhood to adolescence. In addition, these associations seemed not to be phenotype-specific, suggesting potential shared genetic variation across the phenotypes of interest.

## Introduction

Childhood developmental and mental health disorders – encompassing neurodevelopmental, behavioural and emotional disorders – are an emerging global concern, with adverse effects on children’s physical, psychological and social well-being (Scott et al., 2016). Children with neurodevelopmental disorders (NDDs) often exhibit high rates of co-morbid mental health disorders – including internalising symptoms or disorders (e.g., anxiety, fear, sadness, depression, social withdrawal and somatic complaints); and/or externalising symptoms or disorders (e.g., aggression, Attention Deficit Hyperactivity Disorder (ADHD) and conduct disorder (CD)) (Carragher et al., 2015). NDDs and co-morbid mental health disorders in childhood are often also associated with cognitive and emotional regulation impairments (Tajik-Parvinchi et al., 2021) and difficulties in social communication and interactions (Barkley et al., 2006, Feder and Majnemer, 2007). Thus, childhood-onset NDDs and mental health disorders may impede educational and occupational achievement later in life (Masten et al., 2010, Sallis et al., 2019); and have been found to precede a range of adverse mental health sequelae across the lifespan (Cannon et al., 2002).

The aetiology of NDDs and mental health disorders is complex and multifactorial (Sullivan and Geschwind, 2019); evidence suggests that shared, polygenic contributions may exist (Sullivan and Geschwind, 2019). Prior genome-wide association studies (GWAS) have shown that a substantial portion of genetic liability to mental health disorders is conferred by commonly occurring single-nucleotide polymorphisms (SNPs) distributed across the genome (Purcell et al., 2009). However, such GWASs have limited predictive power, as the variants identified typically have small effect sizes (Purcell et al., 2009), and results are often non-specific (i.e. GWASs of mental health disorders are often not unique to a specific disorder) (Wray et al., 2021). Measures such as polygenic risk scores (PRSs) enable the quantification of common risk alleles carried by an individual for a given disorder; and provide an estimate of genetic liability to a trait at the individual level (Purcell et al., 2009). These scores are calculated by summing the risk alleles an individual (from a target group) possesses, weighted by the effect size estimates (obtained from GWAS summary statistics from a discovery cohort) for a particular set of SNPs (Purcell et al., 2009, Choi et al., 2020).

PRSs have been increasingly used in psychiatric research to operationalise genetic risk associations with phenotypes of interest (Wray et al., 2021, Murray et al., 2021). Emerging population-based studies have suggested that PRSs for NDDs and mental health disorders may be associated with behavioural problems in early childhood (Jansen et al., 2020, Krapohl et al., 2016). For example, in their recent study investigating polygenic risk associations for ADHD and autism spectrum disorder (ASD) in an ADHD and ASD clinical population (in 688 children and adolescents versus 943 adult controls), they found that a PRS for adult ADHD was significantly associated with ADHD diagnostic status in children and adolescents (Jansen et al., 2020). Similarly, a PRS for ADHD (in adults) has also been found to associate with attention and/or hyperactivity traits in children (Groen-Blokhuis et al., 2014; Martin et al., 2014); and with general genetic liability toward “broad psychopathology” in childhood (Brikell et al., 2020). There has also been novel work on associations between a PRS for schizophrenia (SCZ) and emotional and behavioural problems in children (Jansen et al., 2018). Further, SCZ PRS has also been associated with general psychopathology (including emotional and behavioural problems) in children (Neumann et al., 2016).

Granted that there is growing interest in delineating associations between PRSs and NDDs/mental health disorders in childhood and adolescence, there is a paucity of such data, particularly when compared to available data in adults (Krapohl et al., 2016, Jansen et al., 2020). Further, to the best of our knowledge, no prior systematic reviews of these studies have been undertaken. Thus, an opportunity exists to delineate the number of studies in this field and the extent of potential associations. To this end, we aimed to systematically review the existing literature on associations between genetic risk (measured by PRS) for NDDs and mental health disorders.

## Materials and Methods

The review was conducted per the Preferred Reporting Items for Systematic Reviews and Meta-analysis (PRISMA) guidelines (Moher et al., 2009); and subsequently registered on the International Prospective Register of Systematic Reviews (PROSPERO CRD42021274965). An electronic literature search was conducted using PubMed, Scopus, PsycINFO and ISI Web of Science. The primary reviewer (L.B Moyakhe) developed the search strategy in consultation with co-authors and a librarian in the Faculty of Health Sciences, University of Cape Town, South Africa. The search terms included “neurodevelopmental disorder,” “polygenic score”,” polygenic risk,” “genetic risk score,” “adolescent”,” child,” “childhood,” “youth,” and “developmental psychopathology.” In addition, the MESH term “Neurodevelopment*” was used in various combinations (and modified as needed for each database). No search restrictions were added. The search commenced on 1 March 2021 and concluded on 1 June 2021. Full details of the search strategy are provided in **Supplementary Table 1**.

### Study selection criteria

Full text published studies evaluating associations between PRS and neurodevelopment (neurodevelopmental outcomes/disorders); and between PRS and mental health (internalising and/or externalising behaviours) in childhood and adolescence were eligible for inclusion. Eligible studies included participants between the ages of two and eighteen years old. Studies of adult populations (older than eighteen); those focusing on neurological disorders or symptoms; animal models or GWASs without a PRS component; and all studies not published in English, were excluded.

### Quality assessment

Quality assessment of the studies was conducted by two independent reviewers (L.B. Moyakhe and M. Mufford) using Q-Genie, a quality assessment tool for genetic studies (Sohani et al., 2016). Q-Genie comprises 11 items assessing studies for bias in the development of the research question, in the ascertainment of comparison groups, and in the classification of the outcome. Potential sources of bias and appropriateness of sample sizes were also assessed. Each item was marked on a 7-point Likert scale – studies were rated as moderate-quality if scoring a total of >32 but ≤40 on Q-Genie; and good-quality if scoring > 40. Complete details of the quality assessment ratings for each included study are provided in **Supplementary Table 2**.

### Data extraction

A data extraction form (adapted from the Cochrane Library’s data collection form) (Lefebvre *et al*., 2011) was developed for this review, (**Supplementary Material, data extraction summary table. xslx file)**. The form was pilot tested using two randomly selected studies that met inclusion criteria; and was refined when additional studies presented new information or data. The information extracted included sample demographics (e.g., age, ancestry); outcome phenotype (e.g., ADHD/ ASD, executive functioning, internalising disorders); outcome measure (e.g., Child Behaviour Checklist); and the reported statistics (e.g., means and standard deviations or odds ratios). The elimination process involved the removal of duplicates and the appraisal of study titles and abstracts to exclude non-relevant studies.

## Results

### Search results

Study selection was undertaken by the first author (L.B. Moyakhe). The initial electronic literature search yielded a total of 740 papers, of which 94 were duplicates, **Figure 1**. Thereafter, 646 papers were screened; and 629 were removed based on title and abstract. The remaining 17 were reviewed as full texts, and nine were subsequently excluded. A total of 8 articles then underwent further assessment, including quality assessment of the studies (i.e., critical appraisal). This process was repeated by the second reviewer (M. Mufford), and any/all discrepancies were discussed by the two reviewers. An additional seven papers were incorporated during this process, thus yielding a total of 15 papers that were reviewed and critically appraised. Thereafter, one study was removed after failing to meet quality assessment standards thus yielding a final total of 14 studies to review.

**Figure 1:**
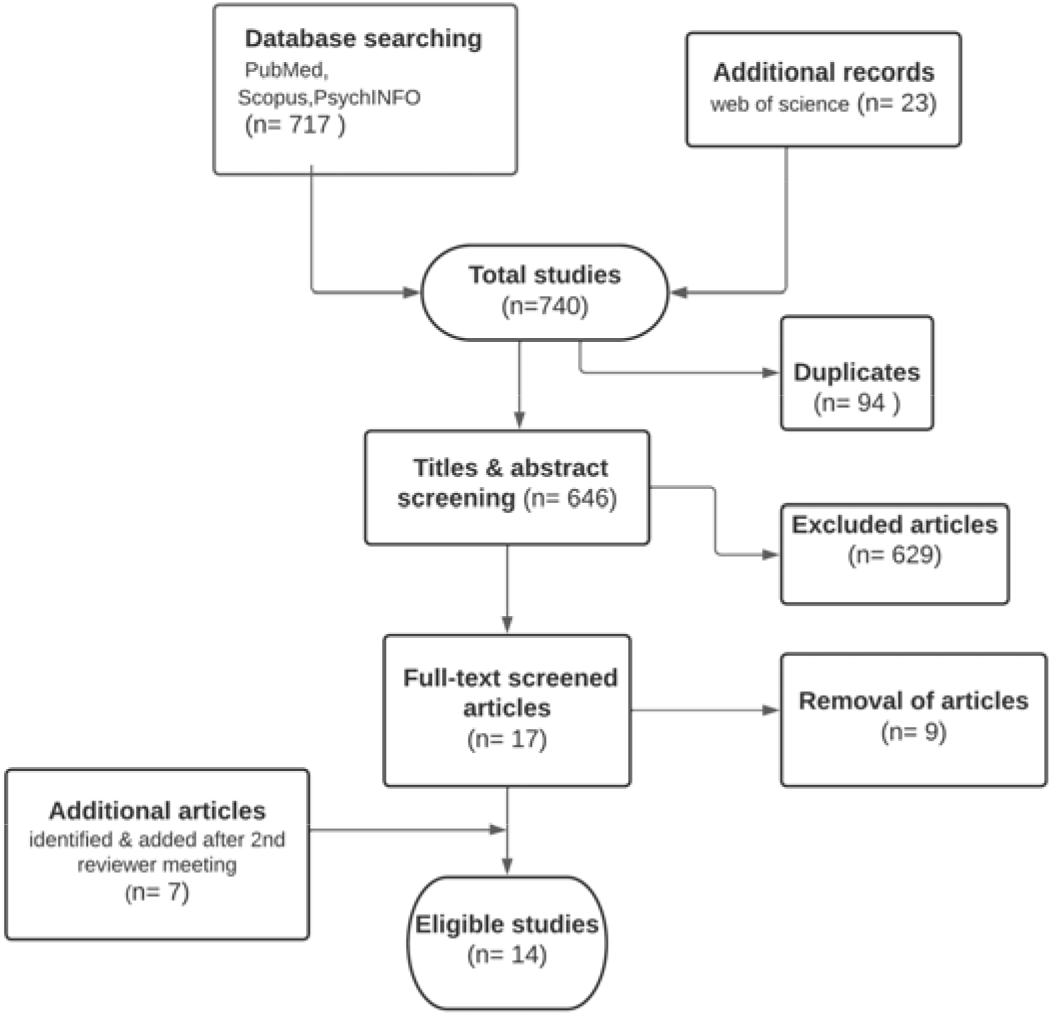
PRISMA flow diagram of study selection.

### Study and sample characteristics

A detailed description of included studies (n=14) is presented in **Tables 1 and 2**. Per inclusion criteria for this review, all study participants were ≤18 years of age. Only data spanning the eligible age(s) were considered in longitudinal and cross-sectional studies. Study participants were primarily of European ancestry; and from well-established cohorts - most commonly the Avon Longitudinal Study of Parents and Children (ALSPAC) (Fraser et al., 2013). Included studies could broadly be categorised as either those investigating (i) PRS and neurodevelopmental outcomes (i.e., language, cognition); or (ii) PRS and mental health outcomes.

### Quality assessment

Quality assessment within and between studies was validated using the Q-genie checklist (Sohani et al., 2016). Studies included in this review were those rated as good quality (i.e., with scores >40). One study was rated as poor quality and subsequently excluded from further analyses; two were rated as moderate quality, and twelve were rated as good quality (**Supplementary Table 2**). Study limitations based on the Q-genie checklist centred mainly on methodological bias, limited sample size and low power. Due to the heterogeneity in the reported outcome measures and methodologies across the studies, a meta-analysis was not feasible in this instance.

### Studies investigating PRS and neurodevelopmental outcomes

Six of the included studies investigated associations between PRS for either neurodevelopmental or mental health disorders, and adverse neurodevelopmental outcomes (i.e., language, cognition, and behaviour) in childhood and adolescence, **Table 1**. The discovery cohorts included the GWAS of British and Irish children with ADHD (Martin et al., 2015); the Psychiatric Genomics Consortium Bipolar Disorder (PGC-BD) working group (Coleman et al., 2020); and the Psychiatric Genomics Consortium Schizophrenia (PGC-SCZ) working group (Ripke et al., 2014). Predictor phenotypes of interest in these cohorts were ASD, ADHD, bipolar disorder (BD), SCZ, and depression (DEP). Target cohorts included ALSPAC and the BRain dEvelopment and Air polluTion ultrafine particles in scHool childrEn (BREATH) project (Mar et al., 2013). The outcomes of interest included working memory, attention performance, cognitive domains, several aspects of IQ (e.g., performance IQ, verbal IQ etc.) and social communication (e.g., pragmatic language).

Across the studies, an association between higher ADHD PRS and adverse outcomes in childhood and adolescence was reported – which persisted with ADHD symptom trajectories. For example, in their study of ADHD symptoms over time (assessed at seven longitudinal time points in participants aged four to seventeen years), Riglin and colleagues (2016) reported symptom persistence in participants with higher ADHD PRS. In line with these findings, Sudre et al. (2020) found worsening ADHD symptoms in participants (aged seven to sixteen years) with higher ADHD PRS (Sudre et al., 2020). Similarly, significant associations between ADHD PRS and ADHD clinical diagnosis, and between ADHD PRS and the combined ADHD/ASD status have been reported in children ages six to eighteen (Jansen et al., 2020). Higher ADHD PRS has also been found to be associated with poorer working memory performance and IQ in childhood (Aguilar-Lacasaña et al., 2020, Martin et al., 2015); and with the development of other neurodevelopmental disorders (i.e., social communication problems; impairment of pragmatic language; conduct problems) in children aged seven to nine years old (Riglin et al., 2016). Additionally, SCZ PRS was reported to associate with ADHD symptoms in children and adolescents ages seven to sixteen years old (Nivard et al., 2017)

Associations between BD PRS and adverse outcomes in childhood were also reported. Specifically, an association was found between higher BD PRS and impaired cognitive function (i.e., poor performance IQ, and processing speed) and poor executive functioning in children aged eight years (Mistry et al., 2019b). A PRS for BD was also found to associate with childhood ADHD (Mistry *et al*. (2019a).

### Studies investigating PRS and mental health outcomes

Eight studies investigated associations between PRS for mental health disorders and mental health outcomes in childhood and adolescence, **Table 2**. The discovery cohorts included the working groups from PGC i.e. the PGC Major Depressive Disorder (PGC-MDD)(Wray et al., 2018) amongst those previously mentioned; the Collaborative Study on the Genetics of Alcoholism (COGA) adult GWAS consortium (Edenberg et al., 2005); and the Netherlands Study of Cognition, Environment and Genes (NESCOG) consortium (Dick et al., 2018). Predictor phenotypes of interest included DEP, MDD, neuroticism (NEU), anxiety (ANX), SCZ and BD (Nivard et al., 2017, Kwong et al., 2021). The relevant target cohorts included ALSPAC (Fraser et al., 2013); the COGA adolescent sample (Edenberg et al., 2005); the Netherlands Twin Registry (NTR) (Ligthart et al., 2019); the “Inside Out” outpatient sample (Jansen et al., 2020); the National Child Development Study (NCDS) (Power and Elliott, 2006); and the Norwegian Mother, Father and Child Cohort (MoBa) (Magnus et al., 2016). Outcomes of interest included ADHD symptoms, internalising symptoms, social problems, depressive symptoms, emotional and behavioural difficulties, and externalising behaviour.

Across the studies, higher PRSs for mental disorders were associated with poorer mental health outcomes in children and adolescents. It was reported that higher PRSs for DEP, MDD and NEU were associated with more severe depressive symptoms in adolescence (Kwong et al., 2021). Further, associations of genetic risk for MDD (indexed by PRS for adults) with psychopathology (i.e., all ADHD symptoms, internalising problems and social problems) in children and adolescents (Akingbuwa et al., 2020) were described. A PRS for MDD was also found to associate with depressive symptom trajectories in the early adolescence class (Rice et al., 2019). Genetic risk for SCZ (as indexed by PRS) associations with adverse mental health outcomes were also described. For example, Nivard and colleagues (2017) reported that genetic risk for SCZ (measured by PRS) was associated with psychiatric disorders in childhood (i.e., ANX, DEP, oppositional defiant disorder (ODD)/CD); and that these associations between the SCZ PRS and internalising disorders increased with age (predominantly for ADHD and ODD/CD) (Nivard et al. 2017). Riglin and colleagues (2018) also found that risk for SCZ (measured by PRS) associated with social and behavioural problems at the age of four years; as well as between SCZ PRS and lower performance IQ, poor outcomes in social understanding and impaired language fluency (at age seven to nine years old). Associations have also been reported between a PRS for SCZ and emotional/behavioural problems in early childhood; as well as with CD, hyperactivity, inattention and ODD in middle childhood (Hannigan et al., 2021). Finally, Salvatore and colleagues (2015) reported that a PRS for externalising behaviour in adults associated with externalising and impulsivity-related behaviour in adolescents (aged twelve to seventeen years).

## Discussion

This systematic review of fourteen eligible studies found a number of associations between PRSs for NDDs/mental disorders and neurodevelopmental and mental health outcomes in childhood and adolescence. Five studies showed significant associations between a higher PRS for ADHD, and poorer neurodevelopmental and behavioural outcomes in children (Martin et al., 2015, Aguilar-Lacasaña et al., 2020). These findings are in line with previous work describing associations between higher ADHD PRS and increased risk for neurodevelopmental, externalising and depressive symptoms in participants aged nine to twelve years (Brikell et al., 2020); and between ADHD PRS and pragmatic language difficulties at ages five and eight (Askeland et al., 2019).

Two studies found associations between a higher BD PRS to associate with poorer executive functioning and processing speed in childhood (Mistry et al., 2019b, Mistry et al., 2019a), which is consistent with prior evidence of associations between BD PRS and deficits in executive functioning (Biederman et al., 2021). Similarly, two studies reported that genetic risk for SCZ (indexed by PRS) associated with adverse mental health outcomes in childhood and adolescence, and this association was found to be significant (Riglin et al., 2017, Nivard et al., 2017). An additional study found that a higher SCZ PRS associated with an increased risk of behavioural symptoms, hyperactivity and inattention in early childhood (Nivard et al., 2017). This is in keeping with previously reported findings of associations between SCZ PRS and emotional recognition (i.e. reduced speed of emotion identification) (Germine et al., 2016).

The remaining four studies reported shared genetic variation (i.e., shared polygenic contributions) within and between NDDs and mental health disorders. For example, associations between BD PRS and childhood ADHD (Mistry et al., 2019a). These cross- and within-traits associations are in line with prior work demonstrating associations between a PRS for several psychiatric disorders and depressive symptoms in adolescents (Kwong et al., 2021); as well as associations between an MDD PRS (in adults) and psychopathology in children and adolescents (Akingbuwa et al., 2020). Similarly, associations between SCZ PRS and depressive symptoms have been described previously. Further, prior cross-sectional and longitudinal studies have reported a higher prevalence of depression in children with ADHD versus controls (Demontis et al., 2019, Copeland et al., 2013, Lee et al., 2013). Further, the genetic overlap observed in this review may also suggest shared biological pathways underlying the neurodevelopmental and mental health outcomes of interest (Solovieff et al., 2013).

There are some noteworthy limitations of the studies in this systematic review. For example, limited sample sizes of the target cohorts which reduce the prediction power of PRS(Chatterjee et al., 2013). Further, the use of parent-rated assessments of neurodevelopmental and mental health outcomes could have led to over- or under-estimation of these phenotypes (Neumann et al., 2016). Several studies included in this review also used summary statistics from adult GWASs (as discovery datasets). In future, age-matched summary statistics could be informative to explore the age-specific effects of NDDs and mental health disorders (Raffington et al., 2020). Finally – though not an inclusion criterion – studies included in this review were of European ancestry populations, with no representation of other ancestral groups (including African ancestries). This is likely due to the limited representation of ancestrally diverse discovery and target datasets in the existing literature (Peterson et al., 2019, Cavazos and Witte, 2020). Thus, improved diversity and inclusivity in such work is needed to improve generalisability, trans-ancestral transferability and translational potential in future (Cavazos and Witte, 2020, Tishkoff et al., 2009, Peterson et al., 2019).

Limitations specific to this review should also be acknowledged. First – though not formally assessed here – publication bias (e.g., skewing towards studies with positive findings) may have indirectly contributed to the studies included. Second, and due to the search strategy employed, it is possible that not all relevant studies were captured and analysed (potentially due to human error during the elimination of non-relevant studies). Third, though considered a priori, a meta-analysis was not undertaken – due to heterogeneity in study design and outcomes measured across studies.

These limitations notwithstanding, this review – which demonstrates associations between ADHD, BD and SCZ PRS and adverse neurodevelopmental and mental health outcomes in childhood and adolescence – highlights the emerging interest in the genomic underpinnings of NDDs and mental health disorders (Lester et al., 2012). Further, the symptom overlap observed between these disorders may be suggestive of shared biological pathways. In future, further work incorporating more ancestrally diverse cohorts and discovery datasets (ideally originating from child/adolescent GWASs), may contribute to an improved understanding of the genetic architecture of development and mental health in childhood and adolescence.

## Supporting information

Data extraction form

## Data Availability

All the information produced in the present study is contained within the manuscript, however, upon reasonable request, additional details can be provided by the authors.

## Acknowledgements

We would like to thank Dr T. Amos and Mrs D. Brey for their expert consultant support received during the development of this systematic review.

