## Supplementary material for "Polygenic risk associations with developmental and mental health outcomes in childhood and adolescence: A systematic review": Table 1 & 2

**Table 1: Characteristics of included studies - PRS and neurodevelopmental outcomes**

| Reference | Discovery Cohort | Target Cohort | Age (years) | Outcome phenotype (scale) | Sample Size | Polygenic Risk Score | Relevant findings |
|---|---|---|---|---|---|---|---|
| **(Aguilar-Lacasaña et al., 2020)** | • ADHD GWAS N= 55 374<br>• ASD GWAS N= 46 351 | Population-based cohort (BREATHE project) | 7 - 11 | ○ Working Memory: n-back task<br>○ Attention performance: n-back task, ANT (computer version)<br>○ ADHD symptoms: DSM-IV | N= 1667 | - ADHD PRS<br>- ASD PRS | - ↑ADHD PRS assoc with ↓working memory performance |
| **(Jansen et al., 2020)** | • Discovery sample ADHD cases N= 20 183, controls N= 35 191<br>• Discovery sample ASD cases N= 18 381, controls N= 27 969<br>• Discovery sample SCZ cases N= 40 675, controls N=64 643<br>• NESCOG | "Inside out" outpatient sample<br>• ADHD<br>• ASD<br>• Controls | 6 - 18 | ○ ADHD symptoms (CBCL 1.5-5)<br>○ ASD symptoms | • ADHD/ASD N= 688<br>• ADHD N= 280<br>• ASD N= 295 | - ADHD PRS<br>- ASD PRS<br>- SCZ PRS | ADHD PRS assoc with :<br>- ADHD symptoms<br>- Combined ADHD /ASD symptoms |
| **(Martin et al., 2015)** | GWAS of British & Irish children with ADHD diagnosis cases N=727 and controls N= 5081 | ALSPAC | 7 - 10 | ○ IQ (WISC-III)<br>○ Verbal working memory: WISC-III digit span task<br>○ Cognitive inhibitory control: Diagnostic Analysis of the Faces subtest, Counting span task (TEACh), Opposite worlds task<br>○ Facial emotional recognition<br>○ ADHD inattentive & impulsive traits: DAWBA<br>○ social communication: SCDC<br>○ pragmatic language scales: CCC | N= 6832 | - ADHD PRS (in children) | ADHD PRS assoc with:<br>- ↓IQ & working memory performance |

| | | | | | | | |
|---|---|---|---|---|---|---|---|
| **(Mistry et al., 2019b)** | PGC working groups:<br>• BD<br>• SCZ<br>• SCZ vs BD GWAS<br>BD cases N= 20 129 and SCZ cases N= 33 426 | ALSPAC | 8 | ○ Cognitive domains : (WISC-III)<br>General intelligence:<br>○ Verbal IQ: (derived from the information, similarities, arithmetic, vocabulary, comprehension, and forward and backward digit span subtests)<br>○ Performance IQ: (derived from the picture completion, picture arrangement, block design, coding and object assembly subtests)<br>○ Total IQ: (sum of PIQ and VIQ)<br>○ Processing speed: (WISC-III)<br>○ Working memory: (Freedom from distractibility score)<br>○ Problem-solving: (WISC-III)<br>○ Executive function: (TEACh)<br>○ Attention: (TEACh)<br>○ Verbal Learning: (CTNWR)<br>○ Emotion recognition: (DANVA) | N= 8230 | - BD PRS | BD PRS assoc with :<br>- ↓executive functioning<br>- ↓processing speed<br>- ↓performance IQ |

**Table 2: Characteristics of included studies – PRS and mental health outcomes**

| Reference | Discovery Cohort | Target Cohort | Age (years) | Outcome phenotype (scale) | Sample Size | Polygenic Risk Score | Relevant findings |
|---|---|---|---|---|---|---|---|
| **Akingbuwa et al., 2020** | GWAS data from 7 cohorts:<br>• ALSPAC<br>• Child and adolescent twin study (Sweden)<br>• Generation R<br>• MoBa<br>• Northern Finland Birth Cohort of 1986<br>• Twins Early Development Study | Meta-analysis | 6 – 17 | ○ ADHD symptoms<br>○ Internalising symptoms<br>○ Social problems<br><br>Scales used:<br>○ SDQ<br>○ CBCL<br>○ CBCL(Youth Self Report)<br>○ SDQ (Conners' Parent Rating Scale)<br>○ Autism-Tics, ADHD and Other Comorbidities Inventory<br>○ Screen for Child Anxiety Repated Emotional Disorders<br>○ Short Mood and Feelings Questionnaire<br>○ Screen for Child Anxiety Related Emotional Disorders<br>○ Short Mood and Feelings Questionnaire<br>○ Rating Scale for Disruptive Behavior Disorders | N= 42 998 | - MDD PRS<br>- NEU PRS | MD PRS assoc with :<br>- Childhood ADHD symptoms,<br>- Internalising problems |
| **Kwong et al., 2021** | PGC working groups:<br>• DEP<br>• MDD<br>• ANX<br>• NEU<br>• SCZ | ALSPAC | 10 - 24 | ○ Depressive symptoms (self-reported) (SMFQ) | N= 6302 | - DEP PRS<br>- MDD PRS<br>- ANX PRS<br>- NEU PRS<br>- SCZ PRS | - ↑ PRS for DEP, MDD, NEU: assoc with adverse DEP symptoms |

| | | | | | | | |
|---|---|---|---|---|---|---|---|
| **(Mistry et al., 2019a)** | PGC-BD | ALSPAC | 7 - 11 | ○ Emotional and Behavioural difficulties: SDQ<br>○ Assessment of childhood ADHD: DAWBA and DSM-IV<br>○ Assessment of borderline personality trait: CI-BPD | N= 8230 | - BD PRS | - BD PRS assoc with ADHD symptoms |
| **(Nivard et al., 2017)** | PGC -SCZ2 | • NTR<br>• ALSPAC | 7 - 15 | NTR:<br>○ Psychopathology: DSM-IV<br>○ Anxiety: CBCL (maternal rating)<br>○ Depression: CBCL<br>○ OCD/ODD: CBCL (maternal rating)<br>ALSPAC:<br>○ Psychopathology: (DAWBA) | • NTR: N=2588<br>• ALSPAC: N= 6127 | - SCZ PRS | - SCZ PRS assoc with childhood psychopathology |
| **(Rice et al., 2019)** | • PGC-MDD<br>• PGC-ADHD<br>• PGC-SCZ | ALSPAC | 10 - 18 | ○ Depressive symptoms trajectories : SMFQ | N= 7543 | - MDD PRS<br>- SCZ PRS<br>- ADHD PRS | - MDD, ADHD and SCZ PRS assoc with DEP symptoms (age 12) |
| **(Salvatore et al., 2015)** | COGA adult GWAS | COGA | 12 - 17 | ○ Subclinical externalising behaviour externalising disorders psychiatric interview<br>○ Impulsivity-related traits: externalising disorders psychiatric interview | N= 248 | - Externalising disorders PRS | - Externalising disorders PRS assoc with subclinical externalising behaviour |

| | | | | | | | |
|---|---|---|---|---|---|---|---|
| **(Riglin et al., 2018)** | • PGC- SCZ<br>• PGC-MDD | NCDS | 7 - 16 | ○ Emotional problems<br>○ Parent report of two depression/anxiety items: Rutter A scale for children (abbreviated version) | N= 5257 | - ADHD PRS<br>- MDD PRS | - SCZ PRS assoc with emotional problems (age 7) |
| **(Hannigan et al., 2021)** | PGC-SCZ | MoBa | 18 months - 8 | ○ Emotional and behavioural psychopathology: (BCL<br>○ Anxiety: SCARED<br>○ Depression: (MFQ<br>○ Disruptive Behaviour Disorders: RS-DBD<br>○ Conduct problems: RS-DBD<br>○ Hyperactivity and inattention: RS-DBD | N= 15 105 | - SCZ PRS | - ↑ SCZ PRS assoc with ↑ behavioural and emotional problems<br>- ↑ symptoms of conduct disorder, ODD, ADHD in middle childhoood |

- **Phenotypes**: DEP=depression, MDD= major depressive disorder, ANX= anxiety, NEU= neuroticism and SCZ= schizophrenia SLI= specific language impairment, ASD= Autism spectrum disorder
- **Measures**: SMFQ= short mood and feelings questionnaire; WISC-III= Wechsler Intelligence Scale - III; TEACh= Test of Everyday Attention for Children; DANVA= Diagnostic Analysis of Nonverbal Accuracy; CI-BPD= the Childhood Interview for DSM-IV Borderline Personality Disorder; SCDC= Social & communication Disorders Checklist; CCC= Children's Communication Checklist, K-SADS Danish version of Schedule for Affective Disorders and Schizophrenia for School-Age Children; TROG-2= Test for Reception of Grammar (Danish version); RIST= Reynolds Intellectual Screening Test. SCARED= Screen for Child Anxiety Related Disorders; RS-DBD= Rating Scale for Disruptive Behaviour Disorders; HRTSE= Hit reaction time; ANT= Attention Network Test; CTNWR= Children's Test of Nonword Repetition
- **Cohorts**: PGC= Psychiatric Genomics Consortium, ALSPAC= Avon Longitudinal Study of Parents and Children; COGA = Collaborative Study on the Genetics of Alcoholism, NCDS= National Child Development Study, SAGE= Study of ADHD, Genes and Environment, MoBa= Norwegian Mother, Father, and Child Cohort, NTR= Netherlands Twin Registry, NESCOG= Netherlands Study of Cognition, Environment and Genes, BREATHE = BRain dEvelopment and Air polluTion ultrafine particles in scHool childrEn
- N's shown only for summary statistics generated by each particular study
- ↑ denotes an increase or higher variable
- ↓ denotes a decrease in a particular variable
