## Supplementary material for "Polygenic risk associations with developmental and mental health outcomes in childhood and adolescence: A systematic review"

### Supplementary Data

Childhood and Adolescents neurodevelopmental disorders and developmental outcomes
Polygenic risk scoring "genetic scoring" or "polygenic scoring"

*Note: Search terms used were broad in order to maintain consistency throughout all databases without being too specific (which results in fewer hits which excludes other vital papers). These broad search terms were intended to also include and capture hits for papers on internalising and externalising behaviours which form part of the crucial inclusion criteria.

Concepts
keywords / Mesh terms
Combine with AND or OR

**Search terms to be used:**

- Developmental psychopathology
- Neurodevelopment disorders (MESH)
- Polygenic risk
- Polygenic score
- Genetic risk
- Child
- Adolescent
- Young people
- Youth

**Supplementary Table 1: Full Search Strategy**

| Database name | MESH | Keywords | Final search | hits |
|---|---|---|---|---|
| **PubMed** | Neurodevelopment disorders | Developmental psychopathology OR Neurodevelopmental Disorders | | |
| | | Polygenic risk OR Polygenic risk score OR Genetic risk score | | |
| | | Child OR Adolescent OR Youth OR Young people | | |

| | | | | |
|---|---|---|---|---|
| | | | **((Neurodevelopmental Disorders[Text Word] OR Developmental psychopathology[Text Word]) AND (Polygenic risk[Text Word] OR Polygenic risk score[Text Word] OR Genetic risk score[Text Word])) AND (Child[Text Word] OR Adolescent[Text Word] OR Youth[Text Word] OR Young people[Text Word])** | 19 |
| **SCOPUS** | | "Development*" "psychopatholog*" OR "Neurodevelopment* Disorder*" | | |
| | | "Polygenic risk*" OR "Polygenic risk score*" OR "Genetic risk score*" | | |
| | | Child* OR Adolescen* OR Youth OR "Young people" | | |
| | | | "Development* psychopatholog*" OR "Neurodevelopment* Disorder*"AND "Polygenic risk*" OR "Polygenic risk score*" OR "Genetic risk score*" AND Child* OR Adolescen* OR Youth OR "Young people" | 674 |
| **Psych Info (via Ebscohost)** | MM (exploded) (S1) "Neurodevelopmental Disorders" | (S2) "Development* psychopatholog*" | | 11,701 |

| | | | | |
|---|---|---|---|---|
| | | OR<br>“Neurodevelopment* Disorder*” | | |
| | | (S3)<br>“Polygenic risk*” OR<br>“Polygenic risk score*” OR<br>“Genetic risk score*” | | 885 |
| | | (S4)<br>Child* OR<br>Adolescen* OR<br>Youth OR<br>“Young people” | | 1,195,759 |
| | | | **S1 AND S2 AND S3 AND S4** | **17** |
| **Web of Science (all databases)** | “Polygenic risk*”<br>“Genetic risk*”<br>“Epigenetic association*”<br>Child*<br>Adolescen*<br>Young people<br>Youth | | | |
| | | | Set 1<br>"polygenic risk *" OR "genetic risk *" OR “Epigenetic association*” | 28 601 |
| | | | Set 2<br>Child* OR adolescen*OR youth OR "young people" | 3 753 903 |
| | | | Set 3<br>"Development* psychopathology " | 2 483 |
| | | | **#1 AND #2 AND #3** | **23** |
| **CINAHL complete (via ebscohost)** | | | | |
| | | (#1)<br>“Development* psychopathlog*”<br>OR | | 11701 |

| | | | | |
|---|---|---|---|---|
| | | "Neurodevelopment* Disorder*" | | |
| | | (#2)<br>"Polygenic risk*" OR<br>"Polygenic risk score*" OR<br>"Genetic risk score*" | | 885 |
| | | (#3)<br>Child* OR<br>Adolescen* OR<br>Youth OR<br>"Young people" | | 1,195,759 |
| | | | **# 1 AND #2 AND #3** | **7** |

**Table 2: Methodological Quality assessment Q-Genie tool**

| Author (year) | Study Rationale | Selection & definition of outcome of interest | Selection & comparability of comparison groups (if applicable) | Technical classification of the exposure | Non-technical classification of the exposure | Other sources of bias | Sample size and power | A priori planning of analyses | Statistical method & control for confounding | Testing of assumptions and inference for genetic analyses | Appropriateness of inferences drawn from results | Q score |
|---|---|---|---|---|---|---|---|---|---|---|---|---|
| Kwong et al 2021 | 4 | 4 | 4 | 7 | 7 | 7 | 3 | 6 | 5 | 6 | 6 | 59 |
| Sudre et al (2020) | 6 | 5 | 5 | 5 | 5 | 3 | 1 | 5 | 4 | 5 | 4 | 48 |
| Hannigan et al (2021) | 5 | 4 | 5 | 5 | 5 | 5 | 3 | 5 | 3 | 3 | 5 | 48 |
| Aguilar-Lacasana et al (2020) | 5 | 5 | 5 | 3 | 5 | 3 | 1 | 5 | 4 | 3 | 5 | 44 |
| Akingbuwa (2020) | 5 | 3 | 5 | 5 | 5 | 5 | 6 | 5 | 5 | 5 | 5 | 54 |
| Jansen (2020) | 5 | 5 | 3 | 5 | 4 | 5 | 2 | 5 | 5 | 5 | 5 | 49 |

| | | | | | | | | | | | |
|---|---|---|---|---|---|---|---|---|---|---|---|
| Martin et al (2015) | 5 | 4 | 5 | 5 | 5 | 5 | 2 | 3 | 3 | 4 | 4 | 45 |
| Mistry et al (a)bipolar | 3 | 4 | 5 | 5 | 5 | 5 | 2 | 3 | 3 | 2 | 3 | 40 |
| Mistry (b) | 5 | 3 | 5 | 3 | 5 | 3 | 2 | 3 | 2 | 2 | 3 | 36 |
| Nivard et al (2017) | 5 | 3 | 5 | 5 | 5 | 5 | 2 | 5 | 5 | 2 | 5 | 47 |
| Nudel et al (2020) | 5 | 5 | 5 | 5 | 5 | 5 | 5 | 2 | 5 | 3 | 7 | 52 |
| Rice et al (2019) | 5 | 3 | 5 | 5 | 5 | 5 | 3 | 5 | 5 | 1 | 5 | 47 |
| Riglin et al (2017) | 5 | 3 | 5 | 3 | 5 | 5 | 3 | 5 | 5 | 1 | 5 | 43 |
| Riglin (2018) | 5 | 2 | 5 | 3 | 1 | 5 | 2 | 5 | 4 | 1 | 5 | 38 |
| Riglin (2016) | 5 | 3 | 5 | 5 | 3 | 3 | 3 | 3 | 3 | 1 | 3 | 40 |
| Salvatore (2015) | 3 | 3 | 5 | 5 | 1 | 4 | 1 | 3 | 3 | 1 | 3 | 33 |

**Poor quality:** 1–2; Good quality: 3–4; **Very good quality:** 5–6; **Excellent quality:** 7. For studies with control groups: scores ≤ 35 indicate poor-quality studies, > 35 and ≤ 45 indicate studies of moderate quality, and > 45 indicate good-quality studies. For studies without control groups: scores ≤ 32 indicate poor-quality studies, > 32 and ≤ 40 indicate studies of moderate quality, and > 40 indicate good-quality studies

NA not applicable
