## Supplementary material for "Polygenic risk associations with developmental and mental health outcomes in childhood and adolescence: A systematic review": Data extraction form

### Q-Genie v1.1 Critical appraisal tool

| | Score (1 to 7) | Kwong et al 2021 |
|---|---|---|
| rationale of study | | 7 |
| Selection & definition of outcome of interest | | 7 |
| selection& comparability of comparison groups | | 4 |
| Technical classification of the exposure | | 4 |
| non-technical classification of the exposure | | N/A |
| Other source of bias | | 5 |
| Sample size and Power | | 6 |
| A priori planning of analyses | | 7 |
| Statistical methods and control for confounding | | 7 |
| Testing: Assumptions & Inferences for genetic analysis | | 7 |
| Appropriateness of inferences drawn from results | | 7 |
| Total scores | | 61 |
| Quality verdict | | good |

| sudre et al 2020 | hannigan et al 2021 | Aguilar-Lacasana et al 2 | Akingbuwa et al 2020 |
|---|---|---|---|
| 6 | 5 | 5 | 5 |
| 5 | 4 | 5 | 3 |
| 5 | 5 | 5 | 5 |
| *5* | *5* | *3* | *5* |
| 5 | 5 | 5 | 5 |
| 3 | 5 | 3 | 5 |
| 1 | 3 | 1 | 6 |
| 5 | 5 | 5 | 5 |
| 4 | 3 | 4 | 5 |
| 5 | 3 | 3 | 5 |
| 4 | 5 | 5 | 5 |
| 48 | 48 | 44 | 54 |
| good | good | good | good |

| Jansen et al 2020 | Martin et al 2015 | Mistry et al 2019 | Mistry et al 2019b |
|---|---|---|---|
| 5 | 5 | 5 | 3 |
| 5 | 4 | 3 | 4 |
| 3 | 5 | 5 | 5 |
| *5* | 5 | 3 | 5 |
| 4 | 5 | 5 | 5 |
| 5 | 5 | 3 | 5 |
| 2 | 2 | 2 | 2 |
| 5 | 3 | 3 | 3 |
| 5 | 3 | 2 | 3 |
| 5 | 4 | 2 | 2 |
| 5 | 4 | 3 | 3 |
| 49 | 45 | 36 | 40 |
| good | good | moderate | good |

| Riglin et al 2016 | Riglin et al 2018 | Salvatore et al 2015 | Nudel et al 2020 |
| --- | --- | --- | --- |
| 5 | 5 | 3 | 5 |
| 3 | 2 | 3 | 5 |
| 5 | 5 | 5 | 5 |
| 5 | 3 | 5 | 5 |
| 3 | 1 | 1 | 5 |
| 3 | 5 | 4 | 5 |
| 3 | 2 | 1 | 2 |
| 3 | 5 | 3 | 5 |
| 3 | 4 | 2 | 5 |
| 1 | 1 | 1 | 3 |
| 3 | 5 | 3 | 7 |
| 37 | 38 | 31 | 52 |
| moderate | moderate | poor | good |

| Rice et al 2019 | Nivard et al 2017 |
| --- | --- |
| 5 | 5 |
| 3 | 3 |
| 5 | 5 |
| 5 | 4 |
| 5 | 5 |
| 5 | 5 |
| 3 | 2 |
| 5 | 5 |
| 5 | 5 |
| 1 | 2 |
| 5 | 5 |
| 47 | 46 |
| good | good |
